# Local emergence and decline of a SARS-CoV-2 variant with mutations L452R and N501Y in the spike protein

**DOI:** 10.1101/2021.04.27.21254849

**Authors:** Jan-Philipp Mallm, Christian Bundschuh, Heeyoung Kim, Niklas Weidner, Simon Steiger, Isabelle Lander, Kathleen Börner, Katharina Bauer, Daniel Hübschmann, Vladimir Benes, Tobias Rausch, Nayara Trevisan Doimo de Azevedo, Anja Telzerow, Katharina Laurence Jost, Sylvia Parthé, Paul Schnitzler, Michael Boutros, Barbara Müller, Ralf Bartenschlager, Hans-Georg Kräusslich, Karsten Rippe

## Abstract

Variants of severe acute respiratory syndrome coronavirus 2 (SARS-CoV-2) are replacing the initial wild-type strain, jeopardizing current efforts to contain the pandemic. Amino acid exchanges in the spike protein are of particular concern as they can render the virus more transmissible or reduce vaccine efficacy. Here, we conducted whole genome sequencing of SARS-CoV-2 positive samples from the Rhine-Neckar district in Germany during January-March 2021. We detected a total of 166 samples positive for a variant with a distinct mutational pattern in the spike gene comprising L18F, L452R, N501Y, A653V, H655Y, D796Y and G1219V with a later gain of A222V. This variant was designated A.27.RN according to its phylogenetic clade classification. It emerged in parallel with the B.1.1.7 variant, increased to >50% of all SARS-CoV-2 variants by week five. Subsequently it decreased to <10% of all variants by calendar week eight when B.1.1.7 had become the dominant strain. Antibodies induced by BNT162b2 vaccination neutralized A.27.RN but with a two-to-threefold reduced efficacy as compared to the wild-type and B.1.1.7 strains. These observations strongly argue for continuous and comprehensive monitoring of SARS-CoV-2 evolution on a population level.

## Introduction

Variants of severe acute respiratory syndrome coronavirus 2 (SARS-CoV-2), the virus causing coronavirus disease 2019 (Covid-19), are of concern with respect to increased viral transmissibility, disease severity, reinfection after naturally acquired immunity, and vaccine efficacy (1). Specifically, variants of concern (VOC) B.1.1.7 (20I/501Y.V1) from the UK (2), B.1.351 (20H/501Y.V2) from South Africa (3), P.1 (20J/501Y.V3) from Brazil (4) and B.1.427/B.1.429 (CAL.20C) from California (5-7) have emerged recently, and the B.1.617 variant from India is currently under investigation with respect to classifying it as a VOC (8, 9). All of the above variants carry mutations in the SARS-CoV-2 spike (S) gene. One common signature mutation of variants B1.1.7, B.1.351 and P.1 is N501Y in the receptor-binding domain (RBD, residues 333–527) (10). This region interacts with the human ACE2 protein (11) and is the primary target of neutralizing antibodies in convalescent and vaccine sera (12). Accordingly, mutations in the RBD are of potential functional relevance for transmissibility and escape from neutralizing antibodies and could reduce vaccine effectiveness (13).

Mutation N501Y has been found to render the virus more infectious, most likely by increasing affinity for the ACE2-receptor (14-17). Other RBD mutations appear to mediate partial escape from neutralizing antibodies. Most notable is the E484K exchange, which is found in VOC B.1.351 and P.1 (14, 15). This mutation was absent in the original UK variant but was recently detected in some B.1.1.7 sequences, indicating further evolution towards immune escape (18).

Other RBD mutations that provide partial resistance to neutralizing antibodies include K417N/T in P.1 and B.1.351 (3, 4, 12) and L452R in B.1.427/1.429 and B.1.617 (5-8, 19). L452R is a marker mutation of the B.1.427/1.429 strain from California (5-7) and is found together with the E484Q mutation in a newly emerging B.1.617 strain from India (8, 20). Outside the RBD, two other S mutations have been associated with a phenotype. The D614G mutation, that occurred early in the pandemic, has been associated with increased transmissibility (21, 22), and the L18F exchange has been linked to faster spreading of SARS-CoV-2 in England (23).

VOC B.1.1.7 has become dominant in the UK in late 2020 accounting for >90% of all infections in 12/2020 and has spread to other regions of Europe and beyond, where it became dominant as well (24). In Germany, B.1.1.7 accounted for the vast majority of all new infections in April 2021 (9). Similarly, VOC P.1 has been responsible for a second wave of infections in Brazil with the epicenter in Manaus (4) and VOC B.1.351 is becoming the major cause of Covid-19 in South Africa (3) with both viruses also spreading globally (1). A study of 2,172 samples collected in California reported that B.1.427/B.1.429 has emerged in late 2020 and increased to >50% of all infections by end of January 2021 with a calculated 18.6-24% increase in transmissibility relative to wild-type (6). B.1.617, another strain with the L452R mutation, has been linked to surge in the number of cases in April 2021 during the second COVID-19 wave in India (20).

Viral genome sequencing is of crucial importance to track the emergence of mutations that define existing and new VOC. By applying a tiling amplicon scheme in combination with multiplex PCR, entire SARS CoV-2 genomes can be sequenced with sufficient coverage in a cost-effective manner (25-27). The comprehensive sequence information obtained by this strategy provides direct insight into the evolution of SARS-CoV-2 genomes and the spreading of variants. In addition, marker mutations identified by sequencing can be exploited to design melting curve-based PCR screens for the rapid detection of relevant variants (28, 29).

Here, we have conducted whole virus genome sequencing of SARS-CoV-2 positive samples from the Rhine-Neckar district in Germany during the first quarter of 2021. We report on the local emergence of a variant designated as A.27.RN that simultaneously carries the N501Y and L452R mutations in the S gene, and its subsequent displacement by variant B.1.1.7. Variant A.27.RN was isolated from patient samples and characterized in tissue culture. Antibody neutralization assays with sera from BNT162b2 vaccinees showed a moderate decrease in neutralizing titers for A.27.RN compared to SARS-CoV-2 wild-type, but better neutralization than for the B.1.351 variant. Our results provide evidence for the transient appearance of a variant of potential concern, carrying the signature N501Y mutation. These observations indicate important differences regarding population spread between N501Y strains that are not recapitulated in infectivity assays in cell culture. Furthermore, they underpin the importance of unbiased surveillance by whole genome sequencing of SARS-CoV-2 positive samples on a population level.

## Materials and methods

### RNA purification

For SARS-CoV-2 PCR and sequencing analysis, RNA was isolated from nasopharyngeal and oropharyngeal swabs applying automated magnetic bead-based nucleic acid extraction protocols. Either the QIASymphony, DSP Virus/Pathogen mini Kit (Qiagen) or Chemagic, Viral DNA/RNA 300 Kit H96 (PerkinElmer) were used, following the manufacturers’ protocols and using the respective devices. The sample input volume was 140 µl, spiked with 10 µl per sample of the internal control (TIB Molbiol) and the elution volume was set to 100 µl. To confirm the mutation pattern after viral passaging in tissue culture, RNA was extracted with the QIAamp Viral RNA body fluid kit which was carried out on the QIAcube system (Qiagen) with manual lysis according to the manufacturer’s protocol.

### RT-qPCR

Samples were screened for SARS-CoV-2 RNA by commercially available dual-target RT-PCR assays using automated analysis for E & orf1a/b employing Cobas 6800 (Roche Diagnostics) or analysis for E & N Gene (TIB Molbiol) or E & S gene (Altona Diagnostics) by Roche LightCycler and LightCycler II. Positive samples with a Ct value ≤35 (reduced to Ct ≤32 in week 10 because of a high rate of inconclusive results in samples with Ct>32) were characterized by VirSNiP SARS-CoV-2 Spike N501Y, DelHV69/70 and K417N Assays (TIB MOLBIOL) using Roche LightCycler II. All commercial assays and devices were used according to the manufacturer’s instructions.

### SARS-CoV-2 genome sequencing

SARS-CoV-2 genomes were sequenced with two approaches, based on the Nextera library generation approach (Cov-seq) and the ARTIC protocol (25) modified and developed by New England Biolabs (NEB, p/n E7658), respectively. For Cov-seq, cDNA was generated with SARS-Cov2 specific primers and a PCR handle in 384-well plates. Reverse transcription (RT) of 2 µl of purified RNA was conducted with the Maxima reverse transcriptase (Thermo Fisher Scientific) and 7.5% PEG-8000. After 50 min at 50 °C and RT inactivation, PCR was performed by adding KAPA PCR mix and custom forward primers and a single reverse primer complementary to the PCR handle. Due to overlapping primer design, the RT and PCR were performed in two separate reactions for each sample. cDNA was purified with SPRI beads at a 0.5x bead ratio and selected samples were measured with Qubit and Tapestation. cDNA was diluted to 0.3 to 0.5 ng/µl and libraries were prepared with the Illumina Nextera kit using a downscaled version with the mosquito system in 384-well plates. The libraries from 384 samples were individually indexed, pooled and purified with SPRI beads.

The ARTIC sequencing generally followed the recommended protocol, but we could further expedite and scale sample processing by implementation of three modifications: (i) With the help of NEB’s technical support team it was possible to omit the magnetic bead clean-up step after PCR amplification of cDNA. (ii) The clean-up of adaptor ligated fragments was conducted on an automated liquid-handling system. (iii) After the final PCR, libraries were pooled, and bead purified with SPRI beads at a 0.9x ratio. All libraries generated were sequenced in pools of up to 384 samples with the Illumina NextSeq 500/550 sequencing instrument in paired-end mode with a read-length of 75 bases from each direction.

### Data analysis

Sequencing data from Cov-seq were processed using the nf-core viralrecon pipeline (30, 31) with the following settings: (i) NC_045512.2 genome, (ii) metagenomic protocol without primer sequence removal, (iii) no duplicate filtering, (iv) minimal coverage of 20 for variant calling, and (v) maximum allele frequency of 0.9 for filtering variant calls. Primer sequences were not removed due to the protocol’s specific amplicon design. Variant calls and reconstructed consensus sequences from iVar were used (32). For the data analysis of the ARTIC sequencing, paired-end sequencing data of read-length 75 bases were filtered for sequencing adapters (trim galore) and host-read contamination (33). Remaining reads were aligned to the NC_045512.2 reference genome using bwa (34). Alignments were sorted and indexed using samtools (35) and quality-controlled using alfred (36). We then masked priming regions with iVar (32). Variant calling employed FreeBayes (37) and bcftools (38) for quality-filtering and normalization of variants. Samtools, bcftools and iVar were also employed for viral consensus sequence generation. Lineage and clade classification of the SARS-CoV-2 sequences was carried out using Pangolin (39) and Nextclade as part of the Nextstrain framework (40).

### Cells and viruses

Vero E6 cells (ATCC#1586) were cultured in Dulbecco’s modified Eagle medium (DMEM, Life Technologies) containing 10% fetal bovine serum, 100 U/mL penicillin, 100 µg/mL streptomycin and 1% non-essential amino acids (complete medium). The original SARS-CoV-2 strain BavPat1/2020 was kindly provided by Christian Drosten, Institute for Virology, Charité Berlin, via the European Virus Archive (Ref-SKU: 026V-03883) and amplified in Vero E6 cells. Cells were inoculated with the original virus isolate (passage 2) at an MOI of 0.01. At 48 h post infection (h.p.i.), the supernatant was harvested, and cell debris was removed by centrifugation at 800xg for 10 min. Virus stocks were produced by one additional amplification in Vero E6 (passage 4), aliquoted and stored at -80 °C. Virus titers were determined by plaque assay as reported earlier (41). To isolate SARS-CoV-2 variants, nasopharyngeal swabs collected from PCR confirmed SARS-CoV-2-positive patients were resuspended and used to inoculate Vero E6 cells that were seeded in 24 well plates at a density of 2.5×10^5^ cells per well. 100 µl of clinical specimen were used to inoculate cells in the first well and five-fold serial dilutions were added to cells in subsequent wells. After 1 h, inoculum was removed and replaced by fresh complete DMEM. Cells were inspected by microscopy for the appearance of a cytopathic effect on a daily basis. In most cases, extensive CPE was observed at four days post-inoculation. Supernatant was collected (passage 0), centrifuged at 1,200 x g for 5 min and 0.35ml were used to inoculate 3×10^6^ Vero E6 cells, seeded into a T25 cm^2^ flask one day prior to inoculation, in a total volume of 5 ml complete DMEM. After two days, CPE was observed, supernatant was collected (passage 1) and used for SARS-CoV-2 genome sequencing. Virus stocks were produced by one further passage in Vero E6 cells (passage 2; B.1.1.7, B.1.351 and A27.RN), virus titer was determined, and stocks were stored at -80 °C.

### Serological analyses

Sera from healthy donors vaccinated with the BTN162b2 SARS-CoV-2 mRNA vaccine were collected at day 10-28 after the second vaccine dose and used immediately or stored at 4°C until use. IgG reactivity against SARS-CoV-2 S1RBD was analyzed by a commercial chemoluminescence immunoassay (Siemens sCOVG; #11207377) run on a Siemens ADVIA Centaur XP instrument according to the manufacturer’s instructions. Interference of sera with RBD-ACE2 interaction was assessed by a titration experiment using serial 1:3 dilutions of sera and a commercial ELISA based test system (GenScript SARS-CoV-2 Surrogate Virus Neutralization Test Kit; #L00847), following the manufacturer’s instructions. Based on the results from these analyses, six sera covering a range of relative reactivities were selected for virus neutralization assays (**Supplementary Table S3**).

### Antibody neutralization tests

Neutralization titers were determined in titration experiments on Vero E6 cells as described previously (42, 43). Vaccinee sera were serially diluted and mixed with the respective SARS-CoV-2 variants. After 1 h at 37 °C, the mixture was added to Vero E6 cells and 24 h later, cells were fixed in the plates with 5% formaldehyde. Virus replication was determined by immunostaining for the viral nucleoprotein using an in-cell ELISA. Values were normalized to the non-treated control (100% infection) and non-infected cells (0% infection). Dose response curves were generated by non-linear regression sigmoidal dose response analysis with Prism 7 (GraphPad Software).

### Determination of viral RNA and infectious virus

Vero E6 cells were seeded into 24 well plates at 2.5×10^5^ cells per well. On the next day, cells were inoculated (MOI=0.05) for 1 h, inoculum was removed, and fresh medium was added. Supernatant was harvested at 48 h post infection. Viral RNA was isolated from supernatants using the NucleoSpin RNA extraction kit (Macherey-Nagel) according to the manufacturer’s specification. Viral RNA quantification was performed using the qScript XLT 1-Step RT-PCR Kit (Quantabio) and probe 2019-nCoV_N1 (5’-FAM-ACC CCG CAT TAC GTT TGG TGG ACC-BHQ1-3’) and the primer set 2019-nCoV_N1 (forward primer: 5’-GAC CCC AAA ATC AGC GAA AT-3’; reverse primer; 5’-TCT GGT TAC TGC CAG TTG AAT CTG-3’). To determine infectious virus titer, 2.5×10^5^ Vero E6 cells were seeded into each well of a 24-well plate. On the next day, cells were inoculated with serial dilutions of SARS-CoV-2 for 1 h. Inoculum was removed, and cells were overlaid with serum free media containing 0.75% carboxymethylcellulose. After 72 h, cells were fixed with 5% formaldehyde for 1 h followed by staining with 1% crystal violet solution. Plaque forming units per ml (PFU/ml) were estimated by counting viral plaques.

### Data availability

SARS-CoV-2 viral genome sequences have been uploaded to the DESH database (https://www.rki.de/DE/Content/InfAZ/N/Neuartiges_Coronavirus/DESH/DESH.html) of the Robert Koch Institute, which will make the data available via the GISAID (https://www.gisaid.org) (44) and the European Nucleotide Archive (ENA, https://www.ebi.ac.uk/ena/browser/home) databases. Representative sequences of A.27.RN with and without the A222V mutation are included as Supplementary Data.

### Ethical statement

This study was approved by the ethics committee of the Medical Faculty at the University of Heidelberg for the analysis of proband samples by whole genome sequencing of the viral RNA (S-316/2021) as well as the virus propagation and neutralization assays and use of sera from vaccinated donors (S-203/2021).

## Results and discussion

### A SARS-CoV-2 variant with L452R and N501Y mutations emerged locally

Due to the recent spreading of VOC in Europe, we implemented whole genome sequencing of all SARS-CoV-2 positive samples detected in the diagnostic laboratory of the Department of Infectious diseases at Heidelberg University Hospital in January 2021. A SARS-CoV-2 variant was identified in 166 out of 1,543 samples acquired from December 2020 to March 2021. According to its phylogenetic lineage A.27, this variant is referred to in the following as A.27.RN (**Fig. 1)**. The first occurrence of A.27.RN in the Rhine-Neckar district was detected in a SARS-CoV-2 positive sample collected in calendar week 1/2021. A.27.RN comprised S gene mutations L18F, L452R, N501Y, A653V, H655Y, D796Y and G1219V in a D614 wild-type background (**Table 1**).

**Table 1.**
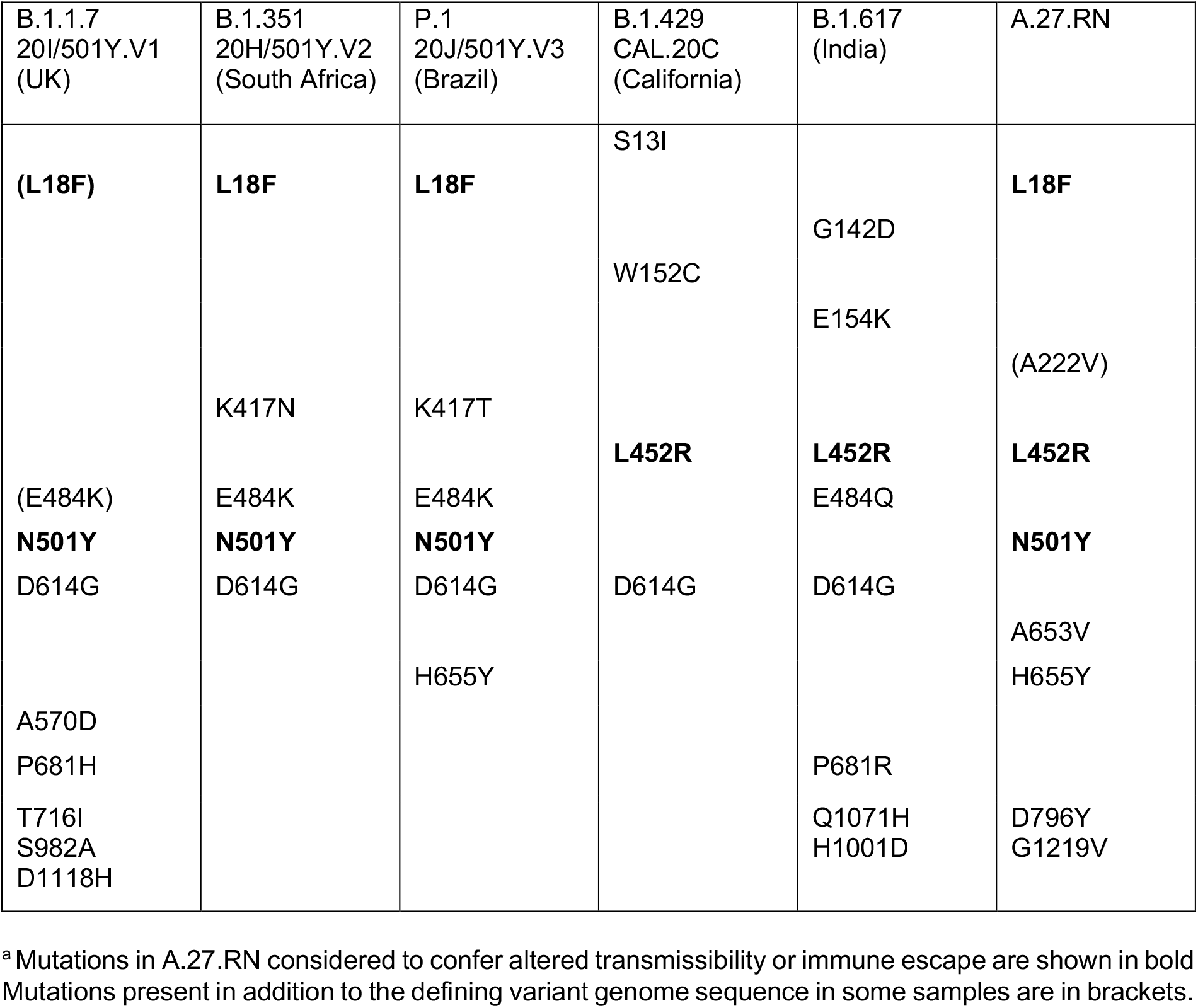
Comparison of S gene mutations in different variants^a^.

**Fig. 1.**
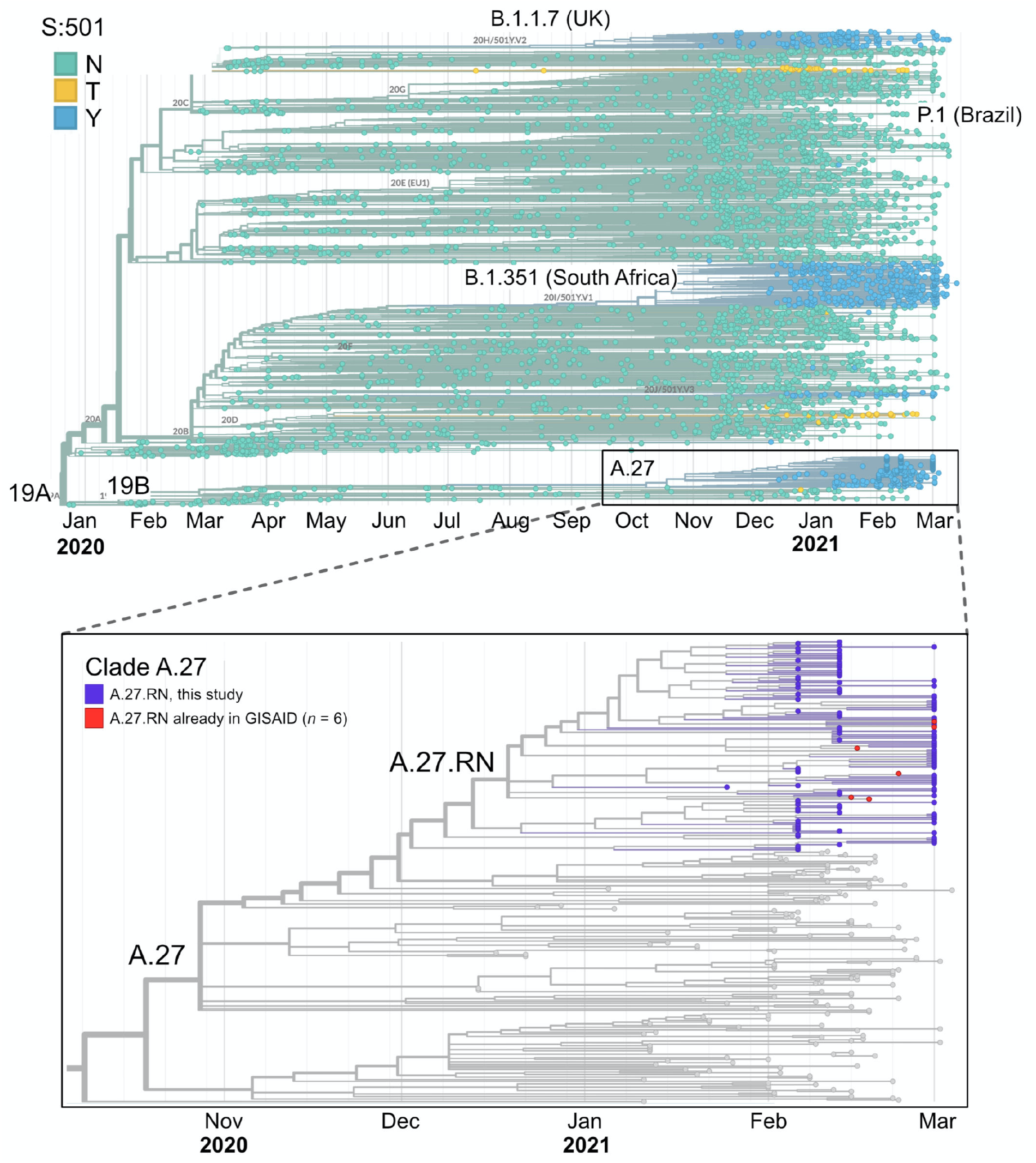
Phylogenetic tree of SARS-CoV-2 variants carrying the N501Y signature mutation. An overview of a global phylogenetic tree analysis is shown in the upper panel. It contains 4,200 genomes sequenced from January 2020 to March 2021 and is colored by the amino acid residue at position 501 of the S protein. Green, N501; yellow, N501T; blue, N501Y. The bottom panel shows a zoom into the A.27 clade with the samples sequenced in our study colored in purple. Six additional samples from GISAID that also fall into the A.27.RN clade are marked in red.

Subsequently, the additional S gene mutation A222V was found in samples collected in February and the relative fraction carrying this additional mutation increased to 10-30% in calendar week 7. The A222V mutation has been described as a marker mutation of the 20A.EU1 strain that has been spreading in Europe in 2020 (45). The location of the mutated residues of A.27.RN in the S protein structure and in the RBD - ACE2 interface is depicted in **Supplementary Fig. S1**. Interestingly, all exchanges in the S protein of A.27.RN are located in unstructured regions at the surface of the protein. In addition to the S gene mutations, A.27.RN carries a D173G mutation and an 8 bp deletion in ORF3a as well as additional mutations listed in **Table S1**. A comparison of A.27.RN with the three main N501Y strains detected so far as well as the B.1.429/CAL.20C and B1.617 strains with L452R is given in **Table 1**. A.27.RN lacks the E484K and the D614G mutations but carries the unusual combination of L452R and N501Y, which has not been observed in the established VOC.

### A.27.RN is phylogenetically separated from other N501Y variants

The sequences obtained were placed into the established SARS-CoV-2 phylogenetic tree (40), yielding the results depicted in **Fig. 1**. A.27 is a newly designated lineage in clade 19B (46). Within this lineage, the A.27.RN sequences form a separate branch (**Fig. 1**) since they carry the unique D173G mutation in ORF3a, which was absent only in 10/166 cases (**Supplementary Fig. 2B**). A.27 is clearly separated from the three currently prevailing N501Y containing strains (**Fig. 1**). The latter, as well as the L452R carrying B.1.427/B.1.429 and B1.617 variants, are classified into clade 20 and lineages therein. At the end of March 2021, 162 A.27 sequences were present in the GISAID database (without the 166 sequences of our study). The total number of A.27 sequences for calendar week 1-11 of 2021 reported to the central database of the Robert Koch Institute for sequencing results in Germany amounts to 424 (47).

**Fig. 2.**
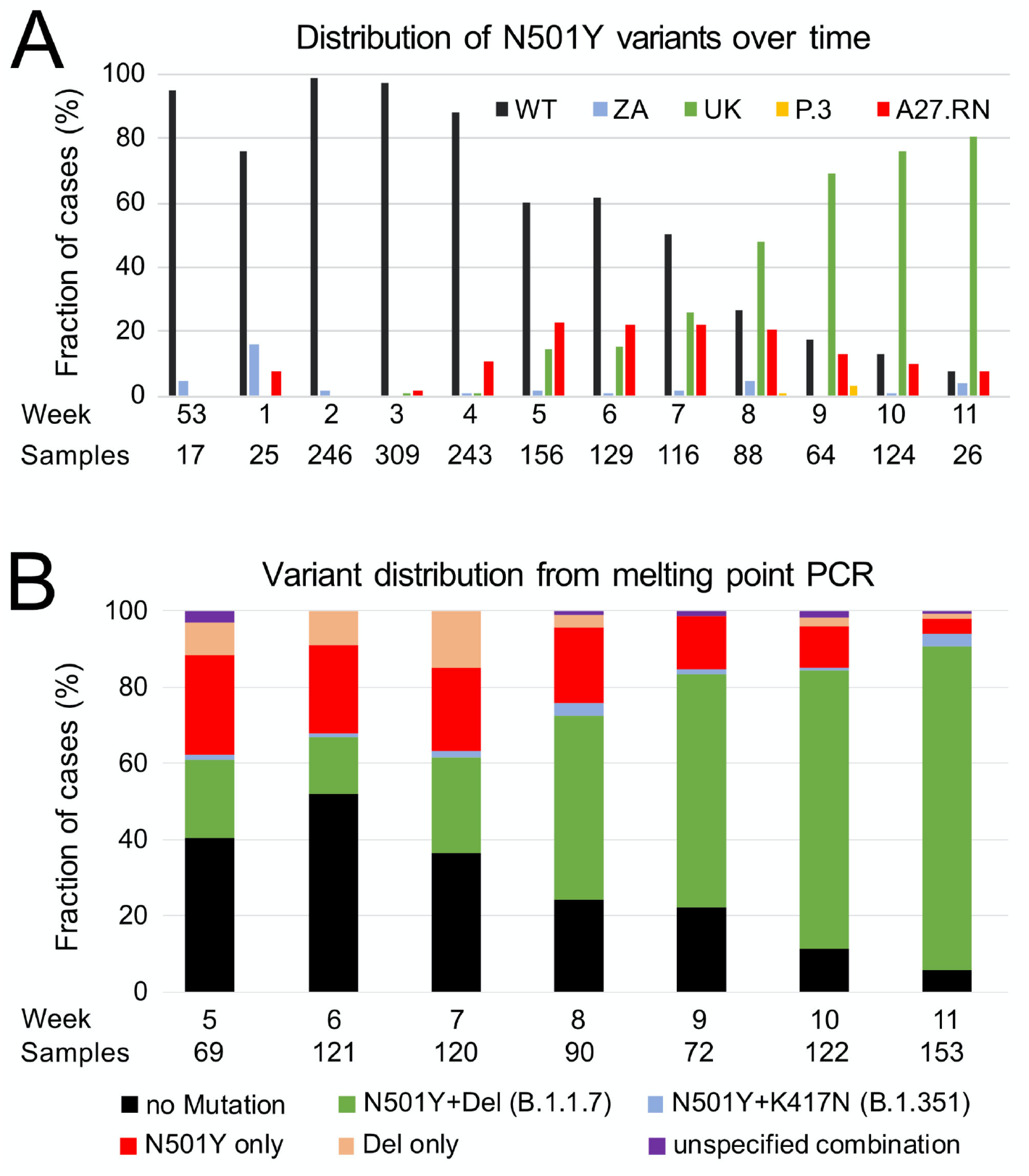
Development of variants in Q1/2021. (**A**) N501Y positive samples split up into the different variants and wild-type virus. Both the fraction of UK B.1.1.7 and the A.27.RN variant increased relatively fast until calendar week seven and started to diverge from week eight. While the ZA variant B.1.351 was present in almost every calendar week, the P.3 mutant occurred very rarely. (**B**) SARS-CoV-2 typing by melting curve analysis over time. Relative amounts of suspected variants are shown for each calendar week confirming the trend seen in the sequencing data. “Del” refers to the HV69/70 deletion.

A.27 sequences in GISAID were previously reported for samples from Denmark, England, Netherlands, Switzerland, Turkey, Slovenia, France, Belgium and Mayotte, an island between northwestern Madagascar and northeastern Mozambique and an overseas department of France (**Supplementary Fig. S2A**). Together with the 166 cases of A.27.RN reported here, three other clustered occurrences of A.27 variants have been reported, namely 12 in Mayotte (January 2021), 92 in France (January to February 2021), and 22 in Slovenia (February 2021), while the remaining 36 were associated with sporadic occurrences. Thus, A.27 strains show a repeated pattern of localized clustered occurrences across different countries during the first quarter of 2021 with 639 cases detected in Germany until calendar week 14/2021 (9).

### Rapid local spread of A.27.RN was followed by outcompetition by B.1.1.7

The first month of 2021 in the Rhine Neckar region was characterized by a rapid local spread of variant A.27.RN. During the period of calendar weeks 53/2020-11/2021 covered in our sample set, N501 wild-type variants were almost completely displaced by variants carrying the N501Y mutation (**Fig. 2A, Table S2**). Initially, A.27.RN dominated and increased to a peak value of ∼58% of all N501Y variants in calendar weeks four and five (**Fig. 2A, Table S2)**. Interestingly, A.27.RN and B.1.1.7 (UK) became the prevailing N501Y variants in calendar week seven with each of them comprising >25% of all samples sequenced (**Fig. 2B**). Subsequently the occurrence of A.27.RN decayed to a 10% fraction in calendar week eleven, while B.1.1.7 became dominant and was found in more than 85% of all samples at that time (**Table S2)**. A.27.RN as well as B.1.1.7 and (in a much lower number of cases) B.1.351 (ZA) were consistently present.

As a faster and less expensive screening readout, we employed Tm-PCR to detect S gene variants N501Y, K417N/K417T and the HV69/70 deletion with data shown here for calendar weeks 5-11 (**Fig. 2C**). Samples were classified as potentially being B.1.1.7, B.1.351, P.1, N501Y only, HV69/70 deletion only or as unknown (**Supplementary Fig. S3A**). Samples for which a valid result could not be obtained for at least one mutation were considered inconclusive. To assess the concordance of the “N501Y only” category with A.27.RN, we matched the Tm-PCR results to sequencing results. From 119 samples categorized as “N501Y only” by Tm-PCR, valid sequencing results could be obtained for 101 samples. Of these, 98 samples (97%) had the A.27.RN sequence (**Supplementary Fig. S3B**,**C**). The remaining three samples (3%) were classified as P.3 (48). The P.3 variant shares the B.1.1.28 ancestor with P.1 and carries E484K but has a distinguishing P681H mutation also present in B.1.1.7. P3 is indistinguishable from A.27 in the Tm-PCR scheme used here, since both variants have N501Y but lack HV69/70 deletion and K417N. Nevertheless, the “N501Y-only” category proved to be a good predictor of the A.27.RN variant in the tested cohort. By exploiting the information obtained from the sequencing data to define a limited subset of mutations, variants of interest can also be identified in a cost-efficient manner with faster turnaround times by the targeted Tm-PCR approach.

### *In vitro* characterization of A.27.RN

To assess replication and spread of A.27.RN in comparison to the BavPat wild-type SARS-CoV-2 strain and the prevailing VOC, variant SARS-CoV-2 A.27.RN, B1.1.7 and B.1.351 were isolated from patient samples and cultured in Vero E6 cells. Culture supernatants of infected cells were prepared and used to determine virus production, which was assessed by quantifying the amount of viral RNA by RT-qPCR and the titer of infectious virus particles using a plaque assay. Overall, ∼3-5 fold lower titers of infectious virus were obtained for all three variants compared to BavPat wild-type strain.

This was reflected by a lower ratio of plaque-forming units to RNA copies for all three variants, arguing for lower specific infectivity of the variant virus particles as compared to the wild-type (**Fig. 3A**). Furthermore, we noted a consistently reduced plaque size in the case of the B.1.1.7 variant compared to the other strains arguing for less efficient spread of B.1.1.7 in this tissue culture system (**Fig. 3B**).

**Fig. 3.**
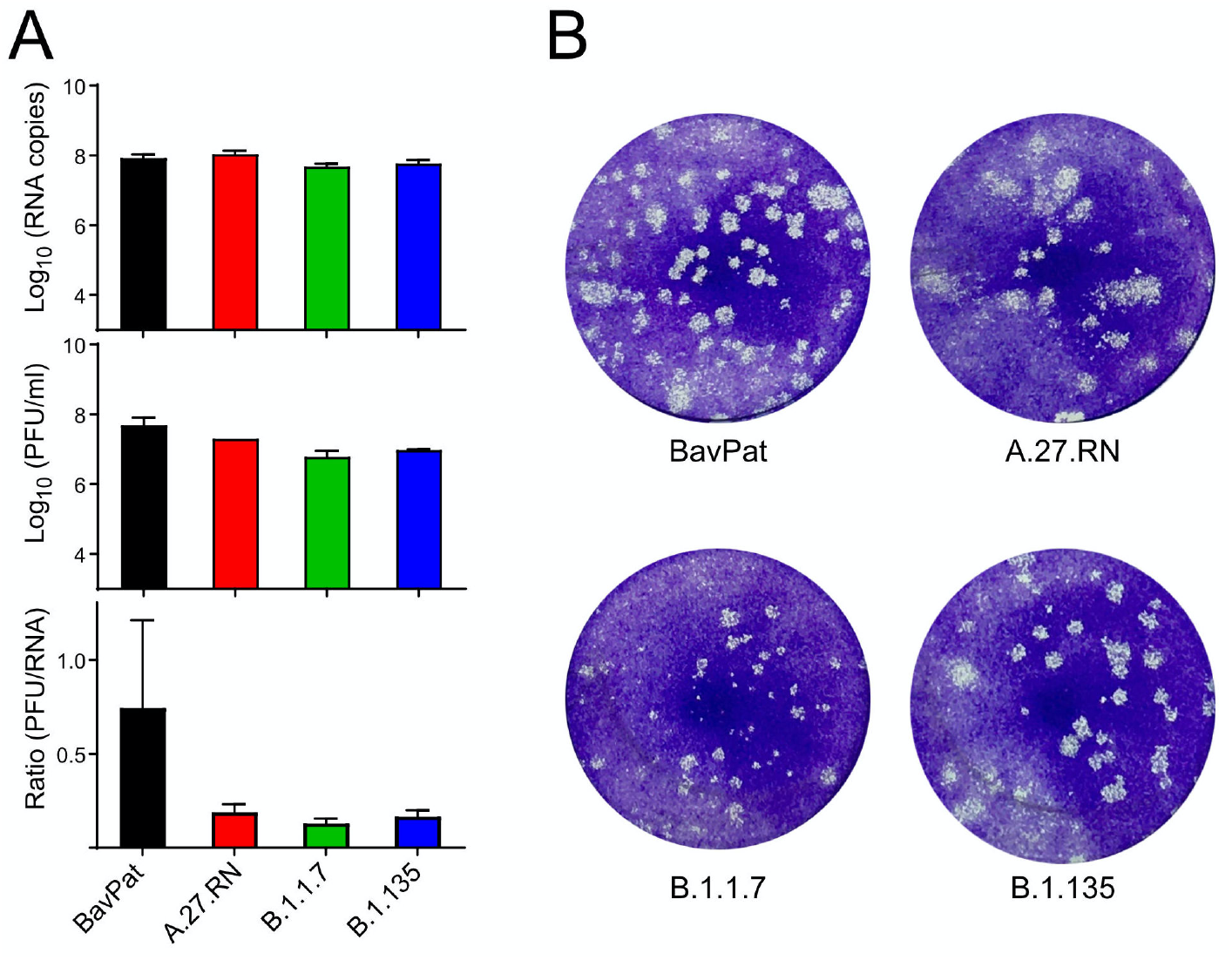
Infectivity titers and RNA-infectivity ratios of different SARS-CoV-2 variants. Vero E6 cells were infected with SARS-CoV-2 wild-type (BavPat) and the indicated variants. (**A**) Quantification of viral RNA and infectivity. Vero E6 cells were infected with given isolates (MOI=0.05) and 48 h later culture supernatants were harvested. Amounts of viral RNA contained in culture supernatants were assessed by RT-qPCR and are given as RNA copies/ml (top panel). The titer of infectious virus was determined by plaque assay and is given as plaque forming units per ml (middle panel). The ratio of RNA copies to PFU (bottom panel) is an approximation for specific infectivity, which is reduced for all 3 variants relative to the wild-type. The graphs show mean values and SD from two biological experiments, each conducted in triplicates. (**B**) Representative images of wells of infected cells, stained with crystal violet at 48 h post infection. Note the smaller plaques obtained with variant B.1.1.7.

### Neutralization of variant A.27.RN is reduced compared to wild-type virus

To evaluate the effect of S gene mutations found in A.27.RN on neutralization capacity of vaccine sera, we performed titration experiments with serum samples from BNT162b2 vaccine recipients. Six samples collected 10-28 days after the second vaccine dose that displayed a range of anti-S1 RBD IgG levels were chosen (**Supplementary Table S3**). Neutralization titers were determined in infection experiments with the A.27.RN isolate and compared against the wild-type control as well as B.1.1.7 and B.1.351 variants (**Fig. 4**).

**Fig. 4.**
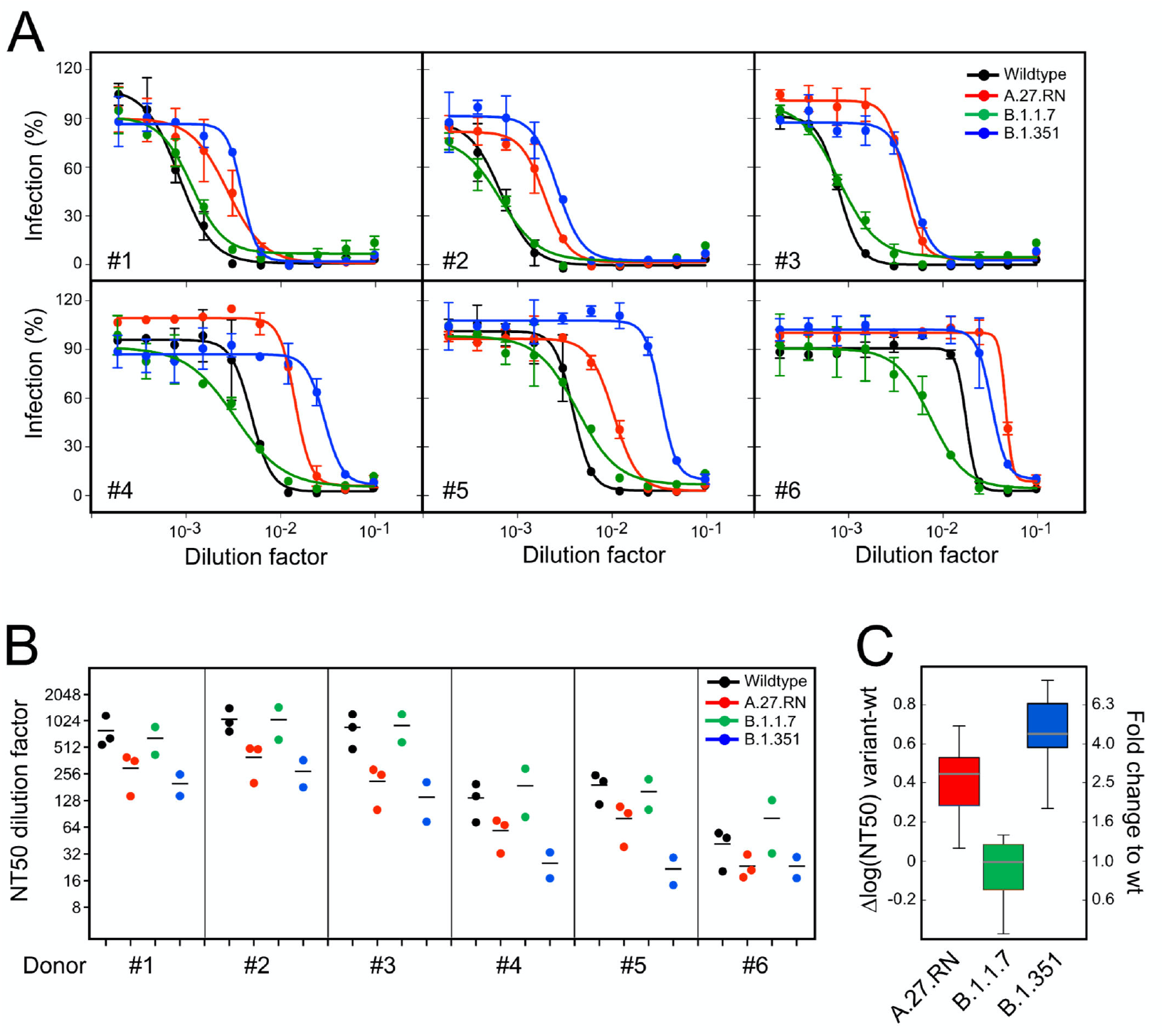
Antibody neutralization capacity of A.27.RN. Neutralizing titers were determined for the sera from six donors vaccinated against SARS-CoV-2 in titration experiments using the indicated SARS-CoV-2 variants on Vero E6 cells. Black: Wild-type (BavPat1/2020); red: A.27.RN; green: B.1.1.7; blue: B.1.351. (**A**) Raw data from one experiment. Data represent mean values and SD from triplicate measurements. The line is a non-linear regression fit to the data that yields the NT50 value, i.e. the dilution at which virus infection is reduced to 50%. (**B**) NT50 dilution factors from three independent experiments. Each data point refers to one independent measurement, lines display mean values. **(C)** Differences of NT50 values of the three variants tested in comparison to wild-type BavPat1/2020.

Neutralizing titers of sera against the wild-type isolate varied from ∼1:40 to 1:1,280, consistent with the different levels of IgG reactivity determined by ELISA. Accordingly, neutralization capacity against the A.27.RN varied somewhat between donors (**Fig. 4A, B**) but was notably reduced compared to wild-type in all cases. Averaged values of the relative difference in NT50 values between variants against wild-type virus yielded a fold-difference and 95% confidence interval (CI) of 2.6 (95% CI = 2.2 to 3.1) for A.27.RN, 0.9 (95% CI = 0.8 to 1.1) for B.1.1.7 and 4.3 (95% CI = 3.1 to 6.0) for B.1.351 (**Fig. 4C**). These values were mostly in line with a recent study that also reports similar serum neutralization values for B.1.1.7 and wild-type but a somewhat higher factor of 9-12 for B.1.351 (12, 49).

## Conclusion

The mutational pattern of A.27.RN is different from other currently prevailing N501Y variants as it combines N501Y and L452R and carries additional S protein and other mutations. According to public databases, the L452R mutation has been acquired by several other independent lineages across multiple countries and continents (5-7, 20, 46). Such repeatedly emerging hot-spot mutations typically indicate strong and probably recent positive selection (17). The spreading profile of A.27.RN on a population level suggests that its transmissibility is higher than that of the SARS-CoV-2 wild-type strain but lower than that of the B.1.1.7 variant. This was not reflected by the growth properties of the virus in tissue culture, where B.1.1.7 exhibited a smaller and A.27.RN a larger plaque phenotype compared to the wild-type BavPat1/2020 reference. Notably, all three variants exhibited a lower specific infectivity, indicating that the observed differences in tissue culture systems may not reflect or predict their actual spread in the population where a variety of other factors come into play.

Our study revealed the regional and clustered appearance of a SARS-CoV-2 variant of potential concern, which rapidly became dominant locally but subsequently was outcompeted by another recently introduced variant on the population level. These results show that the presence of certain signature mutations is not sufficient to predict the spread of different SARS-CoV-2 variants. It can be speculated that the lack of the D614G mutation lowers the transmissibility of A27.RN, since D614G is found in all of the other variants discussed here (**Table 1**) and has been associated with an increased SARS-CoV-2 fitness (21, 22). Vaccine sera were capable of neutralizing A.27.RN, albeit with reduced neutralization titers. This effect was even more pronounced for B.1.351 in line with previous findings (12, 49). Accordingly, it is expected that combinations of the phenotypically relevant L18F, K417N/T, L452R, E484R/Q, N501Y and D614G mutations in the S protein discussed here, as well as newly emerging ones, will become advantageous under immune selection. Continued and comprehensive surveillance of emerging SARS-CoV-2 strains and mutations by unbiased whole genome sequencing in combination with melting point PCR remains therefore of utmost importance for controlling the current pandemic.

## Supporting information

Supplementary Data Set 1. A.27.RN sequence

## Data Availability

SARS-CoV-2 viral genome sequences have been uploaded to the DESH database (https://www.rki.de/DE/Content/InfAZ/N/Neuartiges_Coronavirus/DESH/DESH.html) of the Robert Koch Institute, which will make the data available via the GISAID  (https://www.gisaid.org) database and the European Nucleotide Archive (ENA, https://www.ebi.ac.uk/ena/browser/home). Representative sequences of A.27.RN with and without the A222V mutation are included as Supplementary Data.

## Acknowledgments

We thank Christian Drosten and Barbara Mühlemann from the German National Consultant Laboratory for Coronaviruses for discussion on SARS-CoV-2 variant distribution. We gratefully acknowledge the help of Maria Anders-Össwein with serological analyses and of Marie Bartenschlager with the neutralization assays and thank all diagnostics employees and EMBL GeneCore’s NGS protocol-automation team for their support. This work was supported by the program for surveillance and control of SARS-CoV-2 mutations of the state of Baden-Württemberg, the German Federal Research Network Applied Surveillance and Testing (B-FAST) within the Network University Medicine and the DKFZ@fightCOVID initiative.

## Author contributions

Conceptualization: HGK, KR, RB

Investigation: JPM, CB, HK, NW, KBö, KBa, KLJ, SP, BM, VB, NA, AT

Data Curation: CB, DH

Formal Analysis: SS, IL, TR, JPM, HK, KR Writing – Original Draft Preparation: KR, JPM

Writing – Review & Editing: HGK, KR, JPM, RB, BM, NW, KB, CB, TR, VB, MB

Supervision: HGK, KR, RB, MB

## Supplementary Information

### Supplementary Tables

**Table S1.** A.27.RN mutations outside the S region.

| Table S1. A.27.RN mutations outside the S region |  |
| --- | --- |
| Region | Mutation |
| N | S202N |
| ORF1a | P286L |
| ORF1a | N3651S |
| ORF1b | P1000L |
| ORF3a | V50A |
| ORF3a | D173G |
| ORF8 | L84S |
| ORF3a | del 258/259 (8 bp) |
| ORF8 | del 119/120 (6 bp) |

**Table S2.** Spreading of N501Y variants detected by sequencing^a^.

| Table S2. Spreading of N501Y variants detected by sequencing<sup>a</sup>. |  |  |  |  |  |  |  |  |  |  |  |
| --- | --- | --- | --- | --- | --- | --- | --- | --- | --- | --- | --- |
| Calendar week | Total n | N501Y n | N501Y (%) | UK n | UK (%) | ZA n | ZA (%) | P.3 n | P.3 (%) | A.27.RN n | A.27.RN (%) |
| 53, 2020 | 17 | 1 | 5.9 | 0 | 0 | 1 | 5.9 | 0 | 0 | 0 | 0 |
| 1, 2021 | 25 | 6 | 24 | 0 | 0 | 4 | 16 | 0 | 0 | 2 | 8 |
| 2, 2021 | 246 | 4 | 1.6 | 0 | 0 | 4 | 1.6 | 0 | 0 | 0 | 0 |
| 3, 2021 | 309 | 9 | 2.9 | 3 | 1.0 | 0 | 0 | 0 | 0 | 6 | 1.9 |
| 4, 2021 | 243 | 30 | 12.3 | 2 | 0.8 | 2 | 0.8 | 0 | 0 | 26 | 10.7 |
| 5, 2021 | 156 | 62 | 39.7 | 23 | 14.7 | 3 | 1.9 | 0 | 0 | 36 | 23.1 |
| 6, 2021 | 129 | 50 | 38.8 | 20 | 15.5 | 1 | 0.8 | 0 | 0 | 29 | 22.5 |
| 7, 2021 | 116 | 58 | 50 | 30 | 25.9 | 2 | 1.7 | 0 | 0 | 26 | 22.4 |
| 8, 2021 | 88 | 65 | 73.8 | 42 | 47.7 | 4 | 4.5 | 1 | 1.1 | 18 | 20.4 |
| 9, 2021 | 64 | 53 | 82.8 | 43 | 67.2 | 0 | 0 | 2 | 3.1 | 8 | 12.5 |
| 10, 2021 | 124 | 108 | 87.1 | 94 | 75.8 | 1 | 0.8 | 0 | 0 | 13 | 10.5 |
| 11, 2021 | 26 | 24 | 92.3 | 21 | 80.8 | 1 | 3.8 | 0 | 0 | 2 | 7.7 |
| total | 1543 | 467 |  | 278 |  | 23 |  | 3 |  | 166 |  |

<sup>a</sup> The calendar week when the sample was taken is given. UK, B.1.1.7; ZA, B.1.351; The number of a samples *n* as well as the corresponding percentage fraction of the variants is listed.

**Table S3:** Reactivity of sera used for neutralization assays.

| **Table S3: Reactivity of sera used for neutralization assays** |  |  |
| --- | --- | --- |
| **ID #** | **IgG antiS1RBD (Siemens sCOV CLIA) (ABU)** | **Surrogate neutralization titer (IC50)** |
| 1 | >3270 | 1:1150 |
| 2 | >3270 | 1:740 |
| 3 | >3270 | 1:650 |
| 4 | 3032,38 | 1:145 |
| 5 | 2905,94 | 1:90 |
| 6 | 948,3 | 1:88 |

Sera from healthy donors vaccinated with BNT162b2 were collected at day 10-28 after the second vaccine dose. IgG reactivity was determined by CLIA (Siemens sCOV test kit) as described in Materials and Methods. WHO antibody binding units (ABU) were calculated from the index value obtained using the factor provided by the manufacturer. Interference with the ACE2-RBD interaction was determined by a titration experiment using the GenScript SARS-CoV-2 Surrogate Virus Neutralization Test Kit.

### Supplementary Figures

**Fig. S1.**
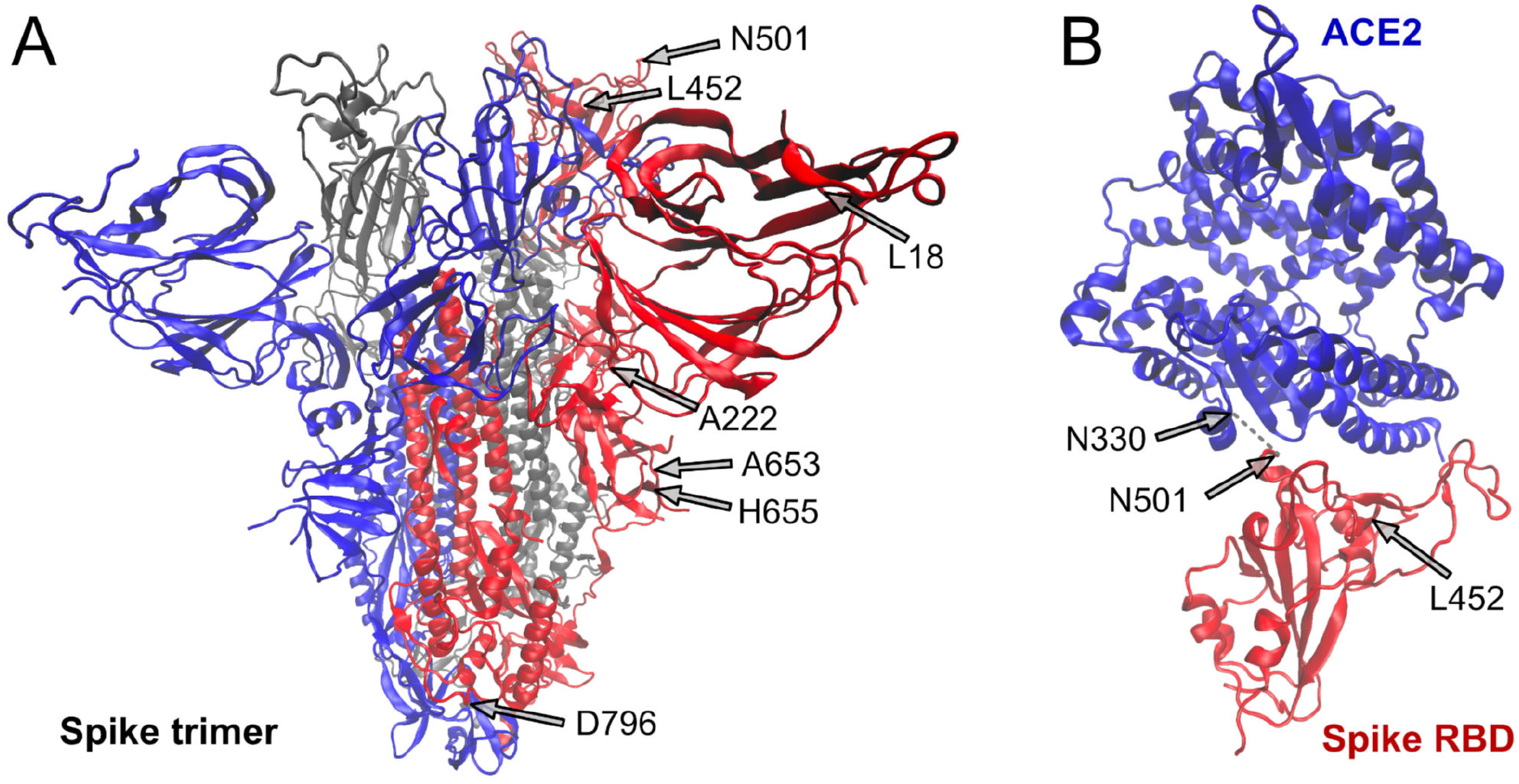
3D structure of SARS-CoV-2 S protein with mutated residues mutated in A.27.RN indicated. A.27.RN mutations were mapped either on the furin cleaved S protein structure using pdb coordinates 6ZGG (50) and on the complex of the S protein RBD and the ACE2 receptor (pdb coordinates 6MOJ) (11) and visualized with VMD viewer (51) (**A**) Complete S protein after furin cleavage. (**B**) Complex of S protein RBD with ACE2 repressor visualization of S protein structure with mutated residues. N501 directly interacts with N330 of the receptor. In contrast, L452 is not in direct contact with ACE2 but creates as hydrophobic patch together with F490 and L492 on the RBD surface.

**Fig. S2.**
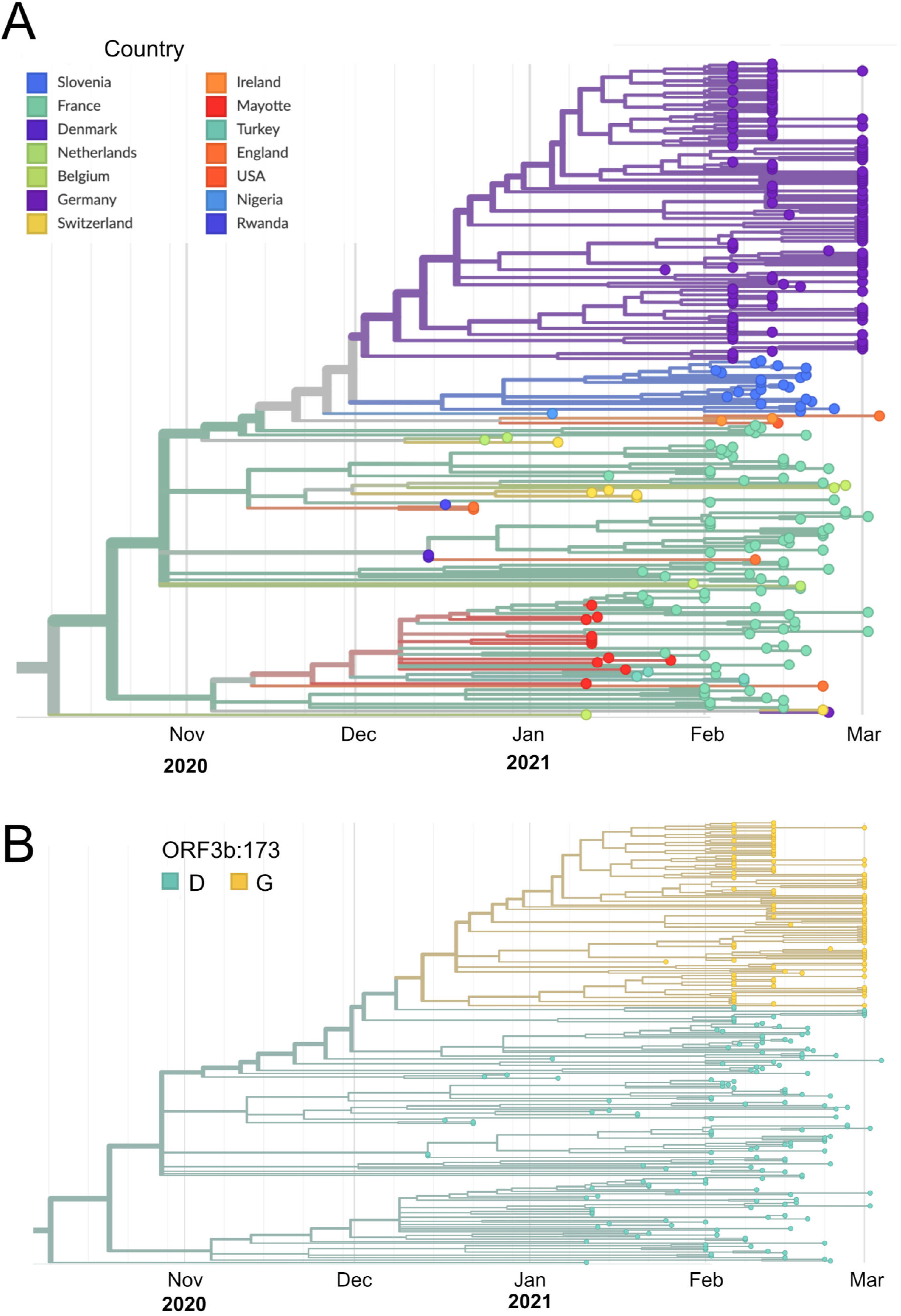
(**A**) Zoom into the A.27 clade as defined by the tool pangolin (39). A total of 162 sequences retrieved from GISAID are shown together with the A27.RN sequences with coloring according to country. (**B**) The same phylogenetic tree as in the bottom panel of Fig. 2 but labeled according to the mutational status ORF3b:173. It can be seen that the A27.RN specific D173G mutation in ORF3b (gold color) separates it from the D173 wild-type background (cyan color) of the other A.27 sequences.

**Fig. S3.**
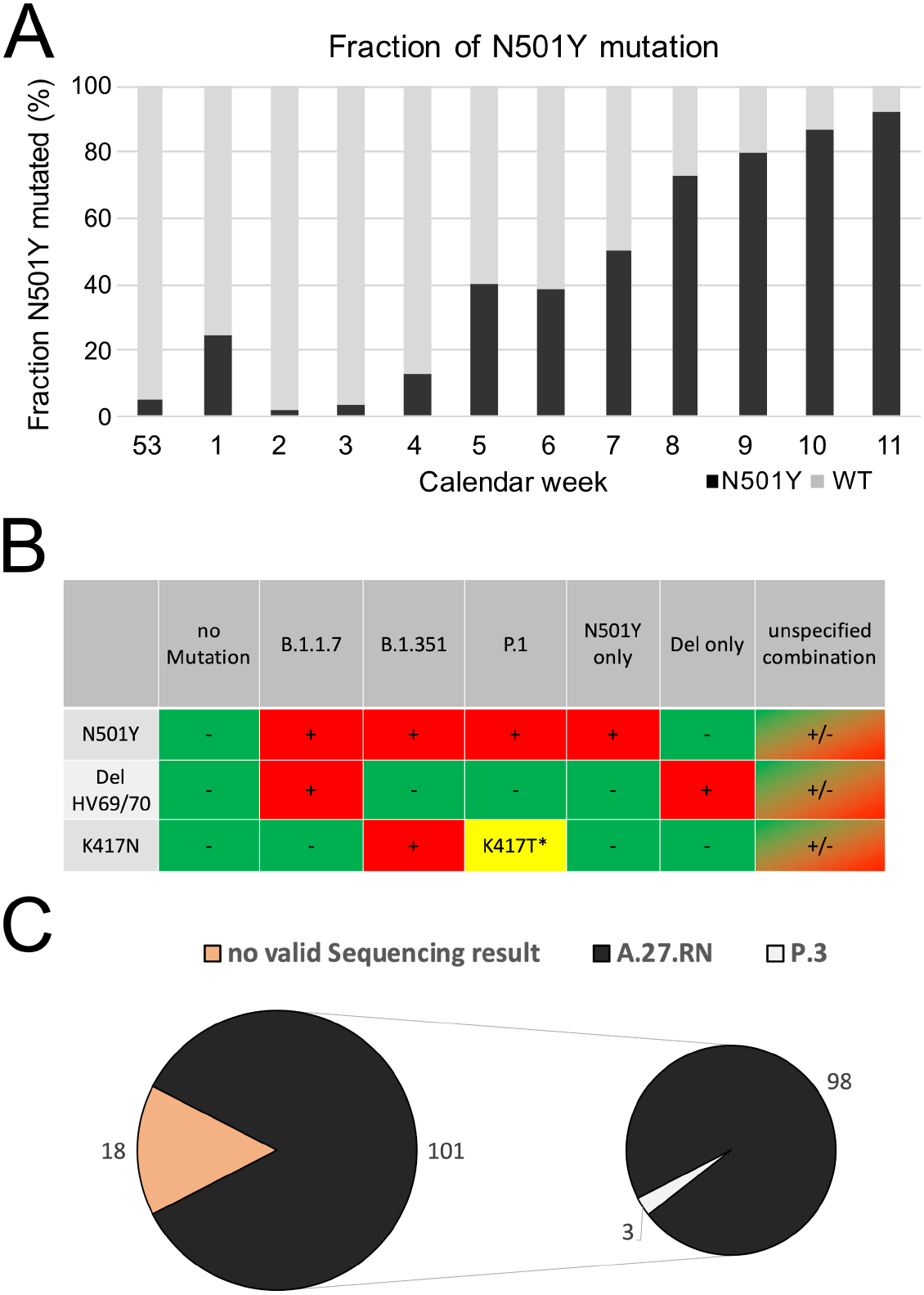
Analysis of SARS-CoV-2 variants with N501Y by sequencing and melting curve analysis. (**A**) Fraction of N501Y mutations present in the samples sequenced. It can be seen that the variants with this mutation have become dominant and comprise 90% of all samples in calendar week 11. (**B**) Samples were analyzed by Tm-PCR for N501Y, K417N and the HV69/70 deletion in the S protein. Preliminary classification was done according to the mutation pattern. K417T is supposed to be represented by a Tm-peak at a lower temperature than 417K (wild-type) in the K417N-assay albeit at reduced specificity. Only samples with valid results in all 3 assays were included. Samples with combinations not shown were classified as “unspecified combination“. (**C**) Comparison of results obtained by sequencings with the Tm-PCR analysis. From 101 matched samples assigned to the “N501Y only” category as a proxy for A.27.RN, 98 samples were indeed this variant, whereas 3 samples were classified as P.3 based on the sequencing analysis.

## Supplementary Data

**Supplementary Data Set 1**

The sequence of the A.27.RN variant as well as its A222V mutated from are provided in FASTA format as a zip file.

